# Prevalence of Anemia of Chronic Disease/Inflammation at a Tertiary Care Hospital in North India

**DOI:** 10.1101/2024.06.26.24309451

**Authors:** Sidheshwar Vishnu Bhendekar, Jay Kirtani, Rahul Naithani

**Author notes:** **Corresponding author** Dr Rahul Naithani, MD, DM, FRCP (Edin), Hematology & Bone Marrow Transplant, Paras Health, Gurugram, Bharat. **Data Availability Statement:** The data used for this research are available from the corresponding author upon reasonable request.

## Abstract

**Background:** There is lack of data from India on prevalence of anemia of chronic disease or inflammation (ACD).

**Patients & methods:** This was a prospective observational cross sectional prevalence study. Anemic patients underwent a complete blood count with peripheral smear, serum ferritin level, iron, total iron binding capacity, transferrin saturation, vitamin B12 and folic acid level, reticulocyte count and stool for occult blood. Other investigations were performed as required according to patient’s clinical profile.

**Results:** Three hundred fifty five patients were enrolled. A total of 109 patients (30.7%) had anemia of chronic disease ACD (30.7%). Sixty three/263 (24%) females had ACD compared to 46/95 (48.4%) males. ACD was four times more common in age group 80 years and above (56.5%) compared to age group 18 to 39 years (13.9%). Seventy two (66%) patients had mild anemia, 19 patients (17%) had moderate anemia and 18 patients (16%) had severe anemia. Diabetes mellitus (44%), hypertension (39%) and chronic kidney disease (25%) were the commonest underlying morbidity. Thirty six patients (33%) had no underlying comorbidity or cause.

**Conclusion:** The prevalence of anemia of chronic disease increases with age. Majority of anemia of chronic disease patients have mild anemia.

## Introduction

Anemia of chronic disease (ACD) or anemia of chronic inflammation (AI) is defined as an anemia associated with a chronic infectious, inflammatory, autoimmune, kidney or neoplastic illness with a characteristic disturbance of iron metabolism.^1^ Acute events such as sepsis, major surgery or trauma can also cause a similar anemia pattern because of acute inflammation and the release of cytokines and hepcidin. These acute medical problems also shorten red cell survival.^2^

ACD/AI is the most common cause of anemia in admitted patients.^3^ It is the second most prevalent cause of anemia after iron deficiency anemia.^4^ It is often confused with iron deficiency anemia and is usually a diagnosis of exclusion. The incidence of ACD increases with age. It is usually observed as a mild to moderate anemia in patients diagnosed with other chronic disease conditions.^5^

ACD is increasingly being viewed as a medical condition that merits direct treatment. It is estimated that up to 40% of all anemias across the globe could be ACD or combined anemias with ACD accounting for more than 1 billion people worldwide.^4,6^ There is lack of data from India on prevalence of ACD. Our search could establish comparative studies with patients of certain diagnoses only. We contribute to scarce data from India by reporting cross-sectional prevalence of ACD in north Indian population with different diagnoses.

## Patients & Methods

This was a prospective observational cross sectional prevalence study. This was part of DNB thesis and study was approved by institute’s ethics committee. The primary objective of this study was to find out the prevalence of anemia of chronic disease. Secondary objective was to identify diseases/conditions associated with the anemia of chronic disease. Anemic patients attending Internal Medicine and Hematology outpatient department (OPD) of a tertiary care hospital were included. Children, ICU patients, patients having known cause of anemia, like iron deficiency, vitamin B12 & folate deficiency, hemolytic anemia and patients with blood loss, like, menorrhagia, hematuria, or gastrointestinal bleeding were excluded. A sample size with a margin of error of 5% on either side and level of confidence 95% sample size was calculated to be 355.

All patients underwent a complete blood count with peripheral smear, serum ferritin level, iron, total iron binding capacity, transferrin saturation, vitamin B12 and folic acid level, reticulocyte count and stool for occult blood. Anemia was defined as hemoglobin (Hb) level < 13 gm/dl in males and Hb < 12 gm/dl in females. Hb <7 g/dl was severe anemia; 7-8.9 g/dl was moderate anemia and Hb 9 and above was defined as mild anemia.^7^ ACD was defined in patients having normochromic, normocytic anemia, low reticulocyte count, low serum iron, normal to low serum transferrin, transferrin saturation <20%, and normal to increased serum ferritin. Other investigations were performed as required according to patient’s clinical profile. These included liver function test, renal function test, serum lactate dehydrogenase (LDH), serum erythropoietin level, antinuclear antibody (ANA) by immunofluorescence, direct coombs test, serum protein electrophoresis and thyroid profile.

## Results

Three hundred fifty five patients were enrolled. A total of 109 patients (30.7%) had anemia of chronic disease ACD (30.7%) was second most prevalent anemia only after iron deficiency anemia (42.2%) (Table 1). Sixty three/263 (24%) females had ACD compared to 46/95 (48.4%) males. The prevalence of anemia of chronic disease increased with age. ACD was four times more common in age group 80 years and above (56.5%) compared to age group 18 to 39 years (13.9%) (Table 2). Seventy two (66%) patients had mild anemia, 19 patients (17%) had moderate anemia and 18 patients (16%) had severe anemia.

**Table 1:** Causes of anemia in 355 patients.

| Type of anemia | Number of patients | Percentage (%) |
| --- | --- | --- |
| Anemia of chronic disease | 109 | 30.7 |
| Iron deficiency anemia | 150 | 42.2 |
| Vitamin B12 deficiency anemia | 19 | 5.3 |
| Hemolytic anemia | 4 | 1.1 |
| Beta thalassemia trait | 9 | 2.5 |
| Iron deficiency anemia + vitamin B12 deficiency | 50 | 14.0 |
| Anemia of chronic disease + vitamin B12 deficiency | 14 | 3.9 |
| Total | 355 | 100 |

**Table 2:** Prevalence of ACD in different age groups.

| Age in years | Number of patients with anemia | Number of patients with ACD | Percentage |
| --- | --- | --- | --- |
| 18 to 39 | 115 | 16 | 13.9% |
| 40 to 59 | 106 | 34 | 32.0% |
| 60 to 79 | 111 | 46 | 41.4% |
| 80 and above | 23 | 13 | 56.5% |
| Total | 355 | 109 | 30.7% |

Other defined causes of anemia were vitamin B12 deficiency, combined deficiency anemias, hemolytic anemia, beta thalassemia minor/trait, or any of above in combination (Table 1). Iron deficiency anemia was the most common cause of anemia in younger as well as older age group. Fourteen patients (3.9%) had pattern of combined ACD and vitamin B12 deficiency. Diabetes mellitus (44%), hypertension (39%) and chronic kidney disease (25%) were the commonest underlying morbidity. Thirty six patients (33%) had no underlying comorbidity or cause (Table 3). Highest prevalent population of ACD without known cause (75%) is present in age group of 18 to 39 years vs 7.6 % in patients age group >80 years.

**Table 3:** Comorbidity/cause in patients with anemia of chronic disease.

| Underlying cause | Number of patients | Percentage (%) |
| --- | --- | --- |
| Diabetes mellitus | 48 | 44.0% |
| Hypertension | 43 | 39.4% |
| Rheumatoid arthritis | 14 | 12.8% |
| Chronic kidney disease | 28 | 25.6% |
| Hypothyroidism | 10 | 9.1% |
| Inflammatory bowel disease | 6 | 5.5% |
| Sarcoidosis | 2 | 1.8% |
| Heart failure | 4 | 3.6% |
| No underlying comorbidity | 36 | 33.0% |

## Discussion

Precise estimates of prevalence of ACD are difficult to ascertain because many patients with anemia are not investigated sufficiently to establish the cause. Moreover, no consensus research criteria exist for the diagnosis of anemia of chronic disease, and patients may have multifactorial causes for anemia, wherein anemia of chronic disease is only a part. Anemia is seen in up to 33-60% patients with rheumatoid arthritis.^8^ Bacterial, viral, parasitic, and fungal infections may all be accompanied by ACD/AI. The prevalence of anemia is likely linked to disease severity.^4^ Malignancy related anemia is seen from 30-60% patients but is usually multifactorial.^9^ In elderly ACD/AI accounts for 35% cases of anemia and is often associated with comorbidities as shown in our study. Chronic disorders such as congestive heart failure, chronic obstructive pulmonary disease (COPD), and excess body weight can also cause ACD.^10^ Patients with chronic kidney disease (CKD) can have ACD and this may also be associated with erythropoietin deficiency. We have earlier shown a prevalence of 37% anemia in patients admitted with Covid infection.^11^ Fifty percent of patients had at least 1 comorbidity which included diabetes mellitus (32%), hypertension (35%), coronary artery disease (8.6%), CKD (5%), COPD (3.9%) and malignancy (1.8%).

ACD is suspected in a patient with an acute or chronic infectious process, inflammatory disorder, or malignant condition who has mild to moderate normocytic, normochromic, hypoproliferative anemia. Typical lab findings are normal to increased serum ferritin, low serum iron level, low transferrin saturation and low total iron binding capacity. Bone marrow aspiration and biopsy are generally not required in the diagnosis of ACD/AI. The hypoferremia associated with ACD is probably caused by defective release of iron from cells, particularly from macrophages, but also from hepatocytes and intestinal epithelium.^12^ Anemia in ACD is usually mild as seen in our study but is common in the elderly population. The anemia of chronic disease contributes hugely to the morbidity of these patients. There is need to identify this disease entity and treat appropriately.

## Conclusion

Anemia of chronic disease is common in patients with systemic diseases. Almost any chronic infection, inflammation, or cancer can cause anemia. Although the second most prevalent after anemia caused by iron deficiency, it is the commonest among patients with chronic illnesses. The prevalence of anemia of chronic disease increases with age. Majority of anemia of chronic disease patients have mild anemia.

## Data Availability

All data produced in the present study are available upon reasonable request to the authors

